# COVID-19 therapeutics for low- and middle-income countries: a review of re-purposed candidate agents with potential for near-term use and impact

**DOI:** 10.1101/2021.03.22.21253621

**Authors:** Daniel Maxwell, Kelly C. Sanders, Oliver Sabot, Ahmad Hachem, Alejandro Llanos-Cuentas, Ally Olotu, Roly Gosling, James B. Cutrell, Michelle S. Hsiang

**Affiliations:** Department of Medicine, University of Texas (UT) Southwestern Medical Center, Dallas, USA; Pandemic Response Initiative, Institute for Global Health Sciences, University of California, San Francisco (UCSF), USA; Department of Pediatrics, Stanford University, Stanford, USA; Institute of Tropical Medicine Alexander von Humbolt (IMTAvH), Universidad Peruana Cayetano Heredia (UPCH), Lima, Peru; Clinical Trials and Interventions Unit, Ifakara Health Institute, Bagamoyo, Tanzania; Department of Pediatrics, UT Southwestern Medical Center, Dallas, USA; Department of Pediatrics, UCSF

**Keywords:** SARS-CoV-2, severe acute respiratory syndrome-coronavirus 2, drug, repositioned, LMIC

## Abstract

Low- and middle-income countries (LMICs) face significant challenges in the control of COVID-19, given limited resources, especially for inpatient care. In a parallel effort to that for vaccines, the identification of therapeutics that have near-term potential to be available and easily administered is critical. Using the United States, European Union, and World Health Organization clinical trial registries, we reviewed COVID-19 therapeutic agents currently under investigation. The search was limited to oral or potentially oral agents, with at least a putative anti-SARS-CoV-2 virus mechanism, and with at least 3 registered trials. We describe the available evidence regarding agents that met these criteria and additionally discuss the need for additional investment by the global scientific community in large well-coordinated trials of accessible agents and their combinations in LMICs. The search yielded 636, 175, and 930 trials, in the US, EU, and WHO trial registers, respectively. These trials covered 17 oral or potentially oral repurposed agents that are currently used as antimicrobials and immunomodulatory therapeutics and therefore have established safety. The available evidence regarding proposed mechanism of actions, clinical efficacy, and potential limitations is summarized. We also identified the need for large well-coordinated trials of accessible agents and their combinations in LMICs. Several repurposed agents have potential to be safe, available, and easily administrable to treat COVID-19. To prevent COVID-19 from becoming a neglected tropical disease, there is critical need for rapid and coordinated effort in their evaluation and the deployment of those found to be efficacious.

## Introduction

The coronavirus disease 2019 (COVID-19) pandemic is having devastating long-term health and socioeconomic impact in low- and middle-income countries (LMICs). Worsening access to essential services, such as immunizations and routine care for preventable and chronic diseases, is also leading to excess all-cause mortality and morbidity^1^. Although lower population density and younger age demographics in some LMICs may be protective against high COVID-19 mortality^2^, poor health infrastructure, including a lack of hospital beds, health care providers and basic supplies, can lead to overwhelmed local health systems with relatively few cases^3^. Effectively responding to COVID-19 outbreaks in LMICs requires the development of new “playbooks” that are contextualized and locally relevant; following the path of high-income countries will most likely lead to wasted resources and ultimately poor epidemic control. Additionally, while vaccine research is moving forward at an unprecedented pace^4, 5^, numerous barriers to effective widespread deployment exist^6,7, 8^. And even with large immunization campaigns, treatments for those who cannot or do not receive the vaccine will still be needed. For many infectious diseases that are vaccine-preventable, antimicrobials remain powerful tools (e.g. bacterial meningitis, varicella, influenza): investment in pharmaceutical interventions should not be compromised in favor of vaccine development.

There is an urgent need for COVID-19 therapeutics that act early in the disease to prevent progression or work as pre- or post-exposure prophylaxis, and others that can treat severely or critically ill patients to prevent mortality. To date, we only have dexamethasone and possibly remdesivir to treat moderate to severe disease, and early non-peer reviewed press reports of improved patient outcomes with injectable monoclonal antibodies for mild illness. However, since the start of the pandemic, researchers have been screening and studying hundreds of available chemicals and drugs with antiviral properties that could be repurposed for treatment of COVID-19. Many of the agents currently in study are used for other diseases and have known safety profiles, which would allow for more rapid scale up of manufacturing at lower cost.

COVID-19 therapeutics must be of course be efficacious, but also accessible for patients in LMICs who may not have easy access to health centers. Oral, transmucosal or transdermal agents will be the easiest to administer at home or an outpatient clinic, followed by intranasal or inhaled formulations through a metered dose inhaler (MDI); these agents are typically less expensive and can be more widely distributed to patients. Subcutaneous, nebulized and intravenous formulations are much less easily administered as they can only be given in the context of a hospital or infusion center, and often require inpatient admission for administration. For example, the current intravenous (IV) formulation of remdesivir has intrinsic barriers to use due to the need for inpatient admission, particularly in countries with large rural populations that live far from health centers^9^.

Here we review current repurposed therapeutics candidates for treatment of COVID-19 currently in advanced clinical trials with potential for easy administration.

## Methods

Using the term “COVID-19” and its associated synonyms (“COVID”, “SARS-CoV-2”, “severe acute respiratory syndrome coronavirus 2”, “2019-nCoV”, “2019 novel coronavirus”, “Wuhan coronavirus”), we searched for trials registered in the United States National Library of Medicine (US NLM, clinicaltrials.gov), European Union Clinical Trials Register (EUCTR, clinicaltrialsregister.eu), and the World Health Organization International Clinical Trials Registry Platform (WHO ICTRP). These results were filtered to exclude non-interventional trials. We then excluded agents with no expected possibility of oral administration, that were rarely studied (n < 3 in WHO ICTRP), or were already part of major guidelines, such as remdesivir.

## Results

The results of the search as of January 14, 2021 are shown in Table 1. The search yielded 636, 175, and 930 trials, in the US, EU, and WHO trial registers, respectively; of these trials, 88, 4, and 72 were completed, respectively. As the WHO ICTRP is a combination of the US and EU and other registers such as the Chinese Clinical Trial Registry (ChiCTR), we present a Preferred Reporting Items for Systematic Reviews and Meta-Analyses (PRISMA) flow diagram for the WHO ICTRP search (figure 1). There were 19 agents, of which 2 were grouped with another structurally similar agents. The final list of 17 oral or potentially oral repurposed agents are currently used as antimicrobials and immunomodulatory therapeutics and therefore have established safety. The agents with >100 trial registered in any of the registers included: hydroxychloroquine, interferon, lopinavir/ritonavir, favipiravir, and ivermectin. Below is a summary of the available evidence regarding each agent’s class, proposed mechanism of action, clinical efficacy, and potential limitations.

**Table 1.** COVID-19 candidate therapeutics with highest potential to be available and easily administrable in the near future.

| Candidate therapeutic | Therapeutic Class | Examples of current uses | Rationale / Mechanism of action <sup>97</sup> | Limitations / Potential obstacles | Trials in United States National Library of Medicine (NLM) clinicaltrials.gov |  | Trials in European Union Clinical Trials Register clinicaltrialsregister.eu |  | Trials in WHO (World Health Organization) International Clinical Trials Registry Platform (ICTRP) list |  |
| --- | --- | --- | --- | --- | --- | --- | --- | --- | --- | --- |
|  |  |  |  |  | Listed | Completed | Listed | Completed | Listed | Phase 4 |
| Hydroxychloroquine<sup>11, 12, 13, 14, 15, 16, 17, 115</sup> | Antimalarial | Malaria, autoimmune conditions, rheumatologic diseases | Block viral entry by impairing terminal glycosylation of ACE2 receptors, proteolytic processing, and endosomal acidification; immunomodulator | Multiple negative trials in all phases of illness, from pre-exposure prophylaxis to treatment of hospitalized patients | 231 | 37 | 70 | 2 | 376 | 32 |
| Azithromycin<sup>18, 19, 20, 21, 22, 23, 24</sup> | Macrolide antibiotic | Community-acquired pneumonia | Alkalinization of endosomal vesicles / lysosomes, potentially inhibiting endocytosis and/or uncoating of enveloped viruses. | No RCTs completed to date except with HCQ/CQ, where it was not shown to be helpful and some signal for harm. | 19 | 1 | 27 | 0 | 32 | 6 |
| Interferon<sup>11, 25, 26, 27, 28, 29, 30</sup> | Biologic response modifier | Hepatitis C, multiple sclerosis | Strengthens IFN response to SARS-CoV-2 infection | Subcutaneous administration (though nasal administration possible), large negative trial | 98 | 9 | 16 | 0 | 99 | 15 |
| Lopinavir/ritonavir<sup>11, 15, 31</sup> | Antiviral | HIV (older regimen) | Protease inhibition | Large negative trial | 73 | 9 | 20 | 0 | 134 | 6 |
| Favipiravir<sup>32, 33, 34, 35</sup> | Antiviral | Investigations for Nipah virus, Ebola, and other flaviviruses | RNA-dependent RNA polymerase inhibition | Safety concerns regarding increases in blood uric acid and teratogenic potential | 42 | 9 | 7 | 0 | 65 | 1 |
| Ivermectin<sup>36, 37, 38, 39, 40</sup> | Antiparasitic | Onchocerciasis, lymphatic filariasis, soil-transmitted helminths, scabies | Nuclear transport inhibition | May be difficult to achieve therapeutic dose | 43 | 12 | 5 | 0 | 70 | 1 |
| Nitazoxanide<sup>41, 42, 43, 44, 45</sup> | Antiparasitic | Cryptosporidiosis, broad-spectrum antiparasitic | Phosphorylation of protein kinase activated by dsRNA; immunomodulator | Trials in early stages | 24 | 3 | 0 | 0 | 27 | 0 |
| Camostat, Nafamostat, and Bromohexine<sup>47, 48</sup> | Serine protease inhibition | Chronic pancreatitis | TMPRSS2 inhibitor | Trials pending | 26 | 0 | 6 | 0 | 32 | 3 |
| Baricitinib<sup>98, 116, 117</sup> | JAK1/JAK2 inhibitor | Rheumatoid arthritis | May inhibit viral entry via clathrin-dependent endocytosis; immunomodulator | IP owned by Eli Lilly | 13 | 2 | 9 | 1 | 16 | 4 |
| Niclosamide<sup>52</sup> | Antiparasitic | Tapeworm infections | Inhibits viral entry by change in endosomal pH; inhibit autophagy | Trials few in number and in early stages | 10 | 0 | 2 | 0 | 10 | 2 |
| Umifenovir<sup>53, 54</sup> | Antiviral | Influenza | Impede trimerization of spike glycoprotein; inhibits adhesion | Trials pending | 10 | 0 | 0 | 0 | 4 | 0 |
| Daclatasvir / Sofosbuvir and Ledipasvir/Sofosbuvir<sup>25, 55, 56, 57, 58, 59, 60, 61, 118</sup> | Antiviral | Hepatitis C | RNA polymerase inhibition | Concerns regarding partial treatment of undiagnosed hepatitis C virus | 7 | 0 | 0 | 0 | 16 | 0 |
| Molnupiravir (formerly MK-4482 and EIDD-2801)<sup>62, 63, 64</sup> | Antiviral | Developed for influenza, investigated for SARS, MERS | Nucleoside analogue | Early phase trials; intellectual property (IP) owned by Ridgeback, possible mutagenicity (disputed) | 5 | 1 | 0 | 0 | 0 | 0 |
| Bemcentinib<sup>65, 66</sup> | AXL kinase inhibition | Trials for malignancy (lung, breast, leukemia, etc) | Block viral entry; enhance IFN | IP owned by BerGenBio | 0 | 0 | 1 | 0 | 2 | 0 |
| Fingolimod<sup>67, 68, 69, 70, 71, 72, 73, 74, 75</sup> | S1P analogue | Relapsing multiple sclerosis | Enhancing S1P can sequester newly generated cytotoxic T cells while preserving memory T cells (immune-modulatory effect). S1P also enhances lung epithelium cell wall integrity | One trial withdrawn | 1 | 0 | 0 | 0 | 2 | 0 |
| Opaganib<sup>76, 77, 78, 79, 80, 81, 83, 119</sup> | ShpK2 inhibitor | Investigated in solid organ tumor, multiple myeloma, and prostate cancer | Inhibiting S1P can have anti-inflammatory and antiviral effects | No solid literature backing postulated claims, no *in vitro* effect on SARS CoV2 | 3 | 0 | 1 | 0 | 4 | 0 |
| Maraviroc<sup>84, 85, 120, 121</sup> | CCR5 antagonist | HIV | potential antagonist to the SARS CoV2 main protease Mpro,3CL pro | MRV had EC50 values above 40 µM (compared with Remdesevir EC50 of 1.1) | 3 | 0 | 1 | 0 | 3 | 0 |
| Doxycycline<sup>89, 90, 91, 93, 122, 123, 124, 125, 126, 127</sup> | S30 bacterial ribosomal inhibitor | Antibiotic | -upregulation of the intracellular zinc finger antiviral protein<br>- an anti-inflammatory profile<br>-could inhibit SARS CoV2 papain-like protease |  | 12 | 4 | 1 | 0 | 15 | 2 |
| Ruxolitinib<sup>94, 95, 96, 97, 128, 129, 130, 131</sup> | JAK1/2 Kinase inhibitor | Used for MF, RA | Potentially inhibits AAK-1, NAK and prevents clathrin-mediated viral endocytosis. Anti-inflammatory agent for cytokine storm | Not as potent AAK1 inhibitor as baricitinib | 16 | 1 | 9 | 1 | 23 | 0 |
| Totals |  |  |  |  | 636 | 88 | 175 | 4 | 930 | 72 |

**Figure 1.**
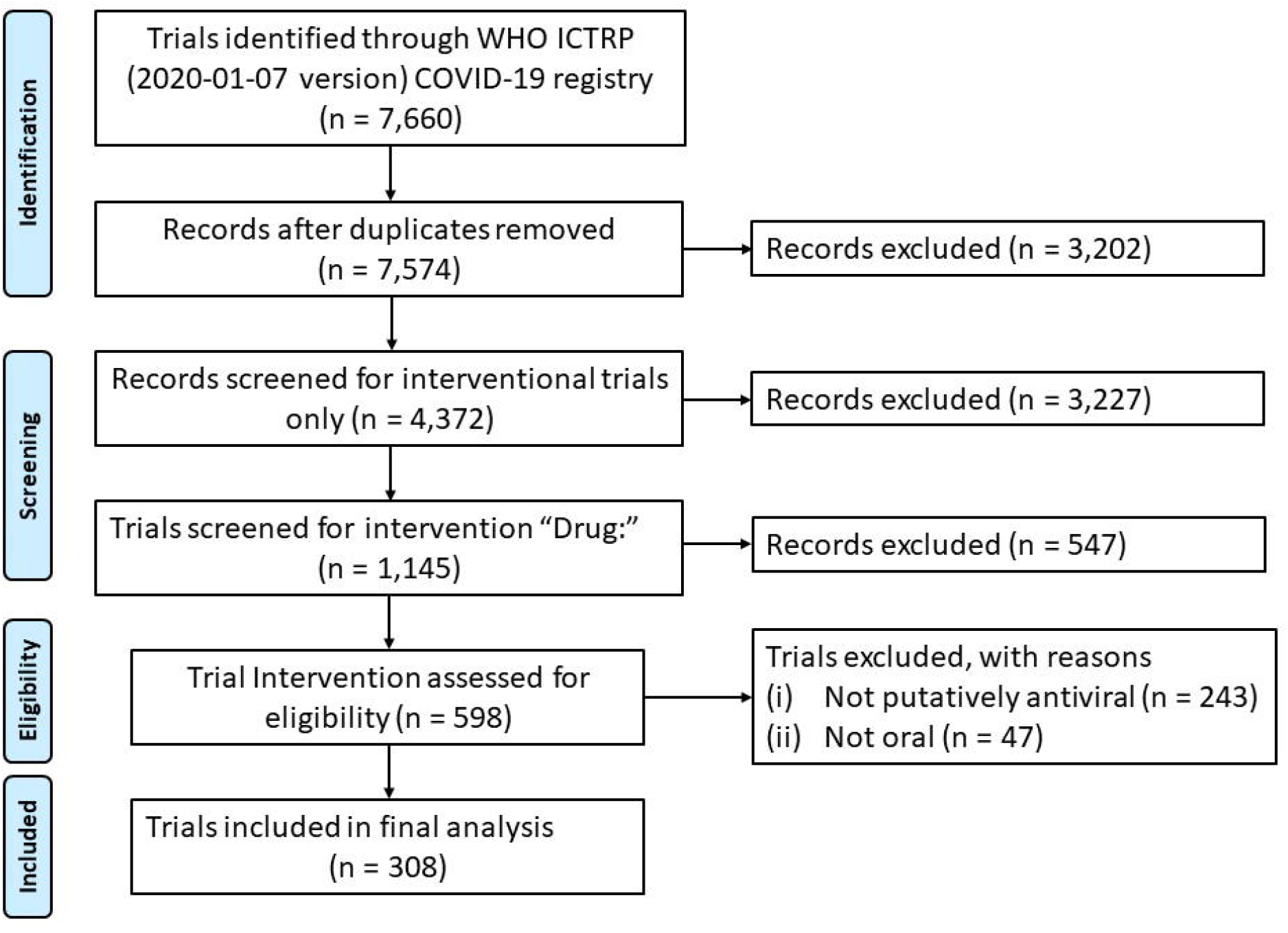
Flow diagram for trial search in World Health Organization International Clinical Trials Registry Platform (WHO ICTRP)

### Hydroxychloroquine (HCQ)

While many trials have been undertaken to investigate the antimalarial and rheumatologic therapeutic CQ and HCQ, a hydroxy derivative of CQ, for treatment of COVID-19, results thus far have been largely negative. Here we focused on HCQ which is less toxic than CQ and has been shown to be more active against SARS-CoV-2 in vitro^10^. The SOLIDARITY trial of the WHO has released interim results as a pre-print and found that none of the 4 treatments tested, including HCQ, reduced 28-day mortality, initiation of ventilation, or duration of hospitalization ^11^. More recently, the RECOVERY Collaborative found no difference in 28-day mortality in 1,561 patients randomized to HCQ or usual care^12^. Of note, median duration of symptoms at time of randomization was 9 days. Considering earlier treatment, again no difference in symptoms found when randomizing 491 patients to early treatment^13^. For prophylaxis, dozens of other trials are underway. One trial of post-exposure prophylaxis in community members and health workers found it to be ineffective ^14^, though there were limitations of this pragmatic trial^15^. Another double-blind, placebo-controlled randomized clinical trial among healthcare workers showed no difference between post exposure prophylaxis of HCQ (600 mg daily for 8 weeks) versus placebo^16^. Another randomized trial of 1483 healthcare workers showing no reduction in incidence of COVID-19 using once or twice weekly HCQ pre-exposure prophylaxis^17^. In summary, all phases of illness have been studied and HCQ has not been found to be effective to date.

### Azithromycin

This macrolide antibiotic has been demonstrated to have a high selectivity index, based on half-maximal inhibitory concentration (IC50) divided by maximal cytotoxic concentration (CC50)^18^. However, in large randomized controlled trials (RCTs) it has been combined with HCQ or CQ and this combination has not shown to be of benefit, and has even shown signals of harm^19, 20, 21, 22^. While no high-quality RCTs of azithromycin alone are available yet, a trial of mass administration to children in Niger resulted in 8 to 14-fold reductions in various coronaviruses based on PCR of the nasopharynx^23^. Further results from at least one large RCT are expected^24^.

### Interferons (particularly beta-1a and beta-1b)

As cytokine mediators that trigger the cellular immune response to viral infections, ^25^ interferons have been used and studied as antiviral therapy for several viral infections^26^. SARS-CoV-2 particularly has been shown to induce particularly low levels of interferon-1^27^. The IC50 of IFN beta-1a against SARS-CoV-2 was found *in vitro* using Vero E6 to increase with time from infection ^28^. However, even as late as 96 hours from infection, high concentrations of IFN beta-1a, in the 50 to 5000 IU/mL range, completely protected Vero E6 cells. Notably, even at 5000 IU/mL, no morphological evidence of cytotoxicity due to IFN-beta-1a used in uninfected cells was noted. Clinically, IFN beta-1a has been evaluated in an open-label, randomized trial of 127 patients comparing LPV/r to triple therapy with LPV/r, IFN beta-1a, and ribavirin^29^. Triple therapy, when administered in the first 7 days, was superior in symptom relief, shortening duration of viral shedding, and shortening duration of hospital stay, all with no difference in adverse events^29^. Subgroup analysis of the trial suggested that IFN beta-1b was the driver of clinical differences between groups. More recently, an Iranian trial of 81 patients comparing the national protocol of HCQ plus LPV/r or atazanavir-ritonavir versus the same protocol plus subcutaneous IFN beta-1a showed no change in time to clinical response (the primary outcome), but did demonstrate decreased 28-day mortality in the IFN beta-1a group (19 versus 43%, p=0.015)^30^. However, the SOLIDARITY trial did not show any benefit in mortality, progression to ventilation, or duration of hospitalization^11^.

### Lopinavir/ritonavir

Lopinavir is a well-studied protease inhibitor used to treat HIV type 1. It is administered in combination with ritonavir, a cytochrome P450 inhibitor, to increase its plasma half-life. As lopinavir/ritonavir (LPV/r) was identified as having *in vitro* activity against SARS-CoV, this agent was used in an open-label clinical trial in a Wuhan hospital (n=199), with participants randomized to LPV/r or standard of care (SOC)^31^. While there was a trend toward lower mortality (19.2% vs 25%), it was not statistically significant. It should be noted that 3 of the 19 patients who died in the experimental arm died less than 24 hours after randomization and never received LPV/r. However, the RECOVERY collaborative found in a large, multicenter randomized trial that this treatment was not associated with reduction in 28-day mortality, duration of hospitalization, or progression to ventilation or death^15^.

### Favipiravir

Favipiravir is another nucleoside analogue shown to have *in vitro* activity against SARS-CoV-2 in Vero cells, with an IC50 of 61.88 µM^32^. Given the CC50 of >400 µM, this yields a SI of > 6.46. While this SI is less favorable than remdesivir’s SI of >129.87, favipiravir was demonstrated to be 100% effective in a post-exposure prophylaxis mouse model after challenge with aerosolized Ebola virus, a virus for which it has a similar IC50^33, 34^. Clinical data on favipiravir for SARS-CoV-2 are still limited. A trial of 80 patients compared favipiravir with interferon alpha against lopinavir/ritonavir with interferon alpha. While the favipiravir group had shorter time to viral clearance (4 vs 11 days), more frequent improvement in chest imaging (91 vs 62%) and fewer adverse events (11% vs 55%), these results are limited by lack of blinding and randomization^32^. An open-label, multicenter phase 3 clinical trial randomized 150 patients with mild to moderate COVID-19 to favipiravir versus standard of care, with a reported faster time to clinical cure and viral clearance by press release although the full data has not yet been published^35^.

### Ivermectin

Given its wide safety margin, ivermectin has been used broadly in mass distribution campaigns to treat illnesses such as onchocerciasis and lymphatic filariasis. More recently, it has been found to limit infection by dengue virus, West Nile Virus, and influenza, with antiviral activity attributed to inhibition of RNA virus interactions with the host nuclear transport proteins^36^. Ivermectin has been tested *in vitro* against SARS-CoV-2, demonstrating reduction in viral RNA in infected cells^36^. However, concentrations needed to achieve viral activity appear to be much higher than what standard doses can achieve in serum. Nonetheless, the drug can concentrate to higher levels in lung ^37, 38^. Possible evidence of this may come from a double-blind, randomized trial of 363 patients with PCR-proven SARS-CoV-2 in Dhaka, Bangladesh. Though not yet in pre-print, the team reported benefit in patients with mild-moderate disease, such as percent of patients experiencing late clinical recovery (23 vs 37%, p<0.03) ^39^. It remains to be seen if this will be born out after peer review and in other trials. Lastly, incorporating ivermectin into COVID-19 treatment could have the added benefit of preventing *Strongyloides* hyperinfection in patients treated with dexamethasone.^40^ As *Strongyloides stercoralis* is hyper-endemic in many LMICs, this could significantly reduce morbidity.

### Nitazoxanide

While not on the WHO List of Essential Medicines, the antiparasitic nitazoxanide has a long history of safety and tolerability since its discovery 30 years ago. It has been investigated as an antiviral previously against MERS in 2016 ^41^, as well as influenza, with variable results^42, 43^. Against SARS-CoV-2, nitazoxanide has a promising ratio of plasma concentration to IC50 for SARS-CoV-2 of 14:1 with standard dosing and could be produced for at a daily cost of $0.10 ^44^. As of January 14, 2021, while 24 clinical trials of nitazoxanide are listed on clinicaltrials.gov, only three have been completed. Notably, the SARITA-2 trial in Brazil, which showed no difference in symptoms at 5 days but did decrease viral load with no serious adverse events noted ^45^.

### Camostat, Nafamostat, and Bromohexine

Camostat and the closely related nafamostat are serine protease inhibitors with decades of clinical use in disseminated intravascular coagulation and pancreatitis, and are well-tolerated^46^. Unlike other agents for SARS-CoV-2, they act at on a human target rather than a viral target called transmembrane serine protease 2 (TMPRSS2)^47^. As this is a necessary agent in the viral spike protein’s function, these agents can partially inhibit SARS-CoV-2 entry into lung epithelial cells. This may be true of other viral infections as well, as an attenuated course of H1N1 influenza was noted in *Tmprss2* knockout mice. While both agents show promising *in vitro* data, nafamostat inhibited SARS-CoV-2 S-mediated entry into host cells with approximately 15-fold-higher efficiency than camostat at a low EC50 ^48^. However, due to the requirement for intravenous administration, nafamostat has been overshadowed by oral camostat in ongoing clinical trials. Lastly, the mucolytic bromohexine, which also inhibits TMPRSS2 protease, was found ineffective in a small (n=18) randomized trial.^49, 50^

### Niclosamide

Niclosamide is another antiparasitic with a well-established safety profile that is currently on the WHO Model List of Essential Medicines ^51^. Although it is used primarily for treatment of cestode infections, niclosamide has been found to have a low IC50 against SARS-CoV-2. Possible mechanistic explanations for this include blocking endocytosis, inhibition of viral replication, and inhibiting receptor mediated endocytosis ^52^. Despite its potential, clinical trials of this agent are very limited.

### Umifenovir

Originally licensed in Russia in 1993 for treatment and prophylaxis of Influenza, umifenovir was found to inhibit SARS-CoV reproduction *in vitro* and clinical trials were quickly undertaken. A retrospective review of 81 hospitalized, non-intensive care unit patients with COVID-19 in China showed no significant differences in outcomes other than longer hospital stays in the umifenovir group^53^. More recently, a meta-analysis of 12 studies, both retrospective and prospective, including a total of 1052 patients, found only a slightly higher rate of PCR-negative testing on hospital day 14 (CI: 1.04 to 1.55)^54^. Given that the end points of the included studies were predominantly symptomatic and laboratory-based with some composite end point, no mortality comparison was made.

### Daclatasvir/sofosbuvir and Ledipasvir/Sofosbuvir

The combination of sofosbuvir and either daclatasvir or ledipasvir has been used to treat hepatitis C and has proven effective for that indication and was well tolerated^55^. More recently, *in silico* studies have been carried out identifying both sofosbuvir and daclatasvir as potential inhibitors of SARS-CoV-2^56, 57, 58^. This combination offer well-established safety and tolerability as combination therapy and has been shown to be safe in patients with renal impairment^25^. Three small trials, sometimes in combination with ribavirin, have been published. All three demonstrated reduced mortality in the treatment group, one achieving statistical significance (p=0.02)^59, 60, 61^.

### Molnupiravir (formerly MK-4482 and EIDD-2801)

Molnupiravir is an orally bioavailable prodrug of beta-D-N4-hydroxycyctidine (NHC) that has demonstrated antiviral activity against Venezuelan Equine Encephalitis Virus^62^. More recently, it has shown *in vitro* activity against SARS-CoV-2 with low IC50s ^63^. When testing for cytotoxicity to establish an estimated therapeutic window, NHC did not cause significant mutagenesis at concentrations as high as 100 µM. Even using the higher IC50 range, this yields a favorable SI of >300 ^63^. Lastly, oral molnupiravir improved lung function in mice infected with MERS-CoV. Currently, it is in phase 2 trials in inpatient and outpatient settings^64^.

### Bemcentinib

Axl is one of a group of tyrosine kinase receptors which has previously been shown to mediate the entry of Zika virus^65^. More recently, an *in silico* screening of approximately 9000 United States Food and Drug Administration (FDA) approved drugs for their potential to interfere with the ability of SARS-CoV-2 to evade innate immune recognition identified the Axl kinase inhibitor bemcentinib as a promising candidate^66^. Moreover, bemcentinib may prevent downregulation of the innate immune response, including IFN production. Lastly, it may inhibit viral cell entry as with Zika virus and others, such as West Nile virus and Ebola virus. Bemcentinib was moved rapidly into the United Kingdom’s ACcelerating COVID-19 dRUG Development (ACCORD) trial and robust clinical data are anticipated.

### Fingolimod and opaganib

The role of modulating Sphingosine 1 phosphate (S1P) pathway either by enhancing or suppressing it is a controversial and sometimes contradictory in terms of the treatment of COVID-19. S1P analogues are being used as part of immune therapy with patients with multiple relapsing multiple sclerosis. Fingolimod is an oral agent that, once phosphorylated, acts as an analogue on S1PR receptors. S1P and its receptor S1P control T cell egress from the thymus and secondary lymphoid organs (SLO) as well. By activating one subtype the S1P receptor (mainly subtype I), naïve and central memory T cells are retained in SLO thus inducing lymphocytopenia^67^. This was proven to be beneficial for patient with autoimmune disease mainly multiple sclerosis where it was proven to reduce relapse rates in clinical trials^68^. The proposed mechanism of the beneficial effect is that Fingolimod traps newly generated encephalitogenic T cells in lymph nodes and prevents migration to the CNS^69^. Even though the physiologic response to viral infections involves lymphocyte migration, Fingolimod does not seem to affect memory B cells and its effect only targets native and central B cells, thus still allowing key immune system responses during therapy. This was proven by several infection models that showed Fingolimod therapy did not affect virus-specific cytotoxic T cells generation^70^. Initial interest to S1P blockade for COVID-19 treatment sprung when several case reports for patients on Fingolimod therapy were diagnosed with COVID-19 and all had very good outcomes^71, 72^. Given that COVID-19-induced ‘cytokine storm’ is a central feature of severe COVID-19, immunomodulatory therapy such as Fingolimod became a subject of interest. Acute lung injury and pulmonary edema which have been extensively described in severe COVID-19 patients are characterized by an increased in vascular permeability. S1P was proven to be a potent angiogenic factor that enhances lung cell wall integrity and an inhibitor of vascular permeability and alveolar flooding in pre-clinical models of acute lung injury^73^. Given that Fingolimod is a S1P analogue, it has been postulated that Fingolimod might also decrease COVID-19-associated increased vascular permeability and associated lung injury^74^. One phase 2 trial was planned to investigate the role of Fingolimod in COVID-19 patients but was later withdrawn. Another point to note is the unclear effect of Fingolimod-induced lymphopenia would have on COVID-19 patients given that lymphopenia is associated with worse outcomes in COVID-19 patients^75^. We could not identify any other trials investigating Fingolimod in COVID-19 patients.

Opaganib is another oral agent that modulates the S1P pathway by inhibiting the enzyme sphingosine kinase isoenzyme 2 ShpK2 thus decreasing levels of S1P. S1P has been shown to be secreted by platelets, monocytes and mast cells in response to inflammation thus promoting inflammatory cascades at the site of injury^76, 77^. In addition, S1P mimics TNF-alpha effect in increasing the expression of cyclooxygenase 2 enzyme and prostaglandin E2^78^. Thus blocking S1P may provide anti-inflammatory properties. That was demonstrated in rodent models of arthritis^79^. Genetic deletion of SPhK2 in mice models conferred protection against Pseudomonas aeruginosa mediated lung inflammation^80^. Although no in-vitro evaluation of antiviral activity of opaganib against the SARS CoV2 virus have been published, there has been previous publications demonstrating antiviral effects of targeting the SphK2 enzyme against the Chikungunya virus^81^, Influenza viruses^82^. Based on the postulated anti-inflammatory and antiviral properties, Opaganib was used on the basis of compassionate use in Israel in 7 patients with reported improvement in both clinical and laboratory parameters^83^. Two trials have since then assessed the role of Opaganib. A phase 2 trial enrolled 40 patients in the US, but was not powered for statistical significance and was focused on efficacy defined by reduction in total oxygen requirement over the course of treatment up to 14 days (NCT04414618). Another global phase 2/3 trial of Opaganib in patients with severe COVID-19 pneumonia (NCT04467840) is currently enrolling patients.

### Maraviroc

Computational bioinformatics have identified chemokine receptor antagonist maraviroc (MRV) as a potential antagonist to the SARS C0V2 main protease M^pro^, 3CL ^pro^, the function of which is regulating replication and transcription by cleaving polyproteins to generate non-structural proteins (NSPs). From a replicase-transcriptase complex. Maraviroc was found to bind to the substrate-binding pocket of M^pro^ forming a significant number of non-covalent interactions^84^. This has not been substantiated with in vitro studies as MRV had EC_50_ values above 40 µM (compared with Remdesevir EC_50_ of 1.1)^85^ thus making MRV an unattractive agent clinically. Nevertheless given the pressing needs created by the pandemic, 3 trials were initiated to investigate the potential benefit of maraviroc for the treatment of COVID-19 patients: 2 are currently recruiting in Barcelona Spain^86^ and Rhode Island Providence Hospital USA^78^. A third trial in Mexico City has maraviroc alone or maraviroc in addition to favipiravir as treatment arms is not yet recruiting.

### Doxycycline

Doxycycline is another potentially repurposed drugs to treat COVID-19 given its antiviral and anti-inflammatory properties. It has showed in vitro activity against SARS-CoV2 infected Vero E cells with median effective concentration EC50 comparable with oral and parenteral formulation ^87^ and has been shown to exhibit antiviral activity against several RNA viruses like dengue virus. The proposed antiviral effect mechanism may be due to upregulation of the intracellular zinc finger antiviral protein which in turns binds to specific viral mRNAs and represses RNA translation, an effect previously demonstrated with Ebola, HIV, Zika and Influenza A viruses ^88, 89^. In addition, doxycycline has been shown to stimulate pro-inflammatory cytokines including interleukin-6 and tumor necrosis factor-alpha^90^ and could inhibit SARS CoV2 papain-like protease ^91, 92^. The Oxford Platform Randomised trial of INterventions against COVID-19 In older peoPLE (PRINCIPLE) which targeted adults with COVID-19 in the outpatient includes an arm for usual care plus doxycycline. In another small scale trial, ivermectin/doxycycline combination vs standard of care was shown to reduce time to recovery and disease progression^93^.

### JAK/STAT inhibitors

In an attempt to mitigate the hyper inflammatory cytokine storm associated with COVID-19, JAK/STAT inhibitors ruxolitinib and baricitinib were both investigated as potential therapeutics. Both inhibit the intracellular pathways of cytokines known to be elevated in severe COVID-19. Baricitinib was also identified as a numb-associated kinase ^25^, with a particularly high affinity for AP-2 associated protein Kinase-1, an important regulator of clathrin-mediated viral endocytosis, and thus could potentially inhibit intracellular viral replication^94, 95^. Several cases series have demonstrated a beneficial effect of baricitinib in treatment of COVID-19 patients^96, 97^. The International Society for Influenza and other Respiratory Virus Disease Antiviral Group Conference reported that the addition of baricitinib to remdesivir in patients hospitalized with COVID-19 was associated with shorter hospital stay (8 vs 7 days, p=0.04) and that there was a decrease in death, though not achieving statistical significance, for death at 29 days in the baricitinib group (5.1 vs 7.8%, p=0.09)^98^. The ACTT-2 trial (Adaptive COVID-19 Treatment Trial-2) was a multinational, multicenter placebo-controlled double randomized trial that demonstrated the combination therapy of remdesivir and baricitinib vs remdesivir alone provided a lower time to recovery in patients requiring high-flow oxygen ^9^. Another JAK1/2 inhibitor, ruxolitinib, was studied in a small randomized trial in Wuhan, China, patients treated with ruxolitinib did not have statistically significant reductions in time to clinical improvement nor survival ^32^. An unpublished Phase III multicenter, randomized, double blind placebo-controlled trial comparing ruxolitinib with standards of care (RUXCOVID) failed to meet its primary endpoint of severe complications ^99^.

## Discussion

This review of candidate COVID-19 therapeutics highlights some of the extensive work already completed in screening individual compounds. It should be noted that the therapeutics arm of the Access to COVID-19 Tools Accelerator, with funding from the Bill and Melinda Gates Foundation, the Wellcome Trust, the Mastercard Impact Fund, National Institutes of Health, and others, have worked to advance this agenda substantially, coordinating the sharing of preclinical compound libraries and clinical research^100, 101^. Another example of such work is the ‘Accelerating COVID-19 Therapeutic Interventions and Vaccines’ or ‘ACTIV’ public-private consortium, involving United States’ government agencies, the European Medicines Agency, and private industry^102^. More critical work remains to be added to the achievements of this team and others toward the design and implementation of adaptive, pragmatic clinical trials specifically targeted for the resource-constrained settings of LMICs. Of note, given the lessons learned from treatments for other viruses such as HIV and hepatitis C virus, it is possible that combination therapy could prove more beneficial than monotherapy alone. The following framework, adapted from other proposed frameworks^103^, has an emphasis on rapid discovery of agents for LMICs. It highlights the aim of advancing candidate agents from pre-clinical screening of compounds to clinical trials, with an intentional focus on the unmet needs in LMICs (fig 2).

**Figure 2.**
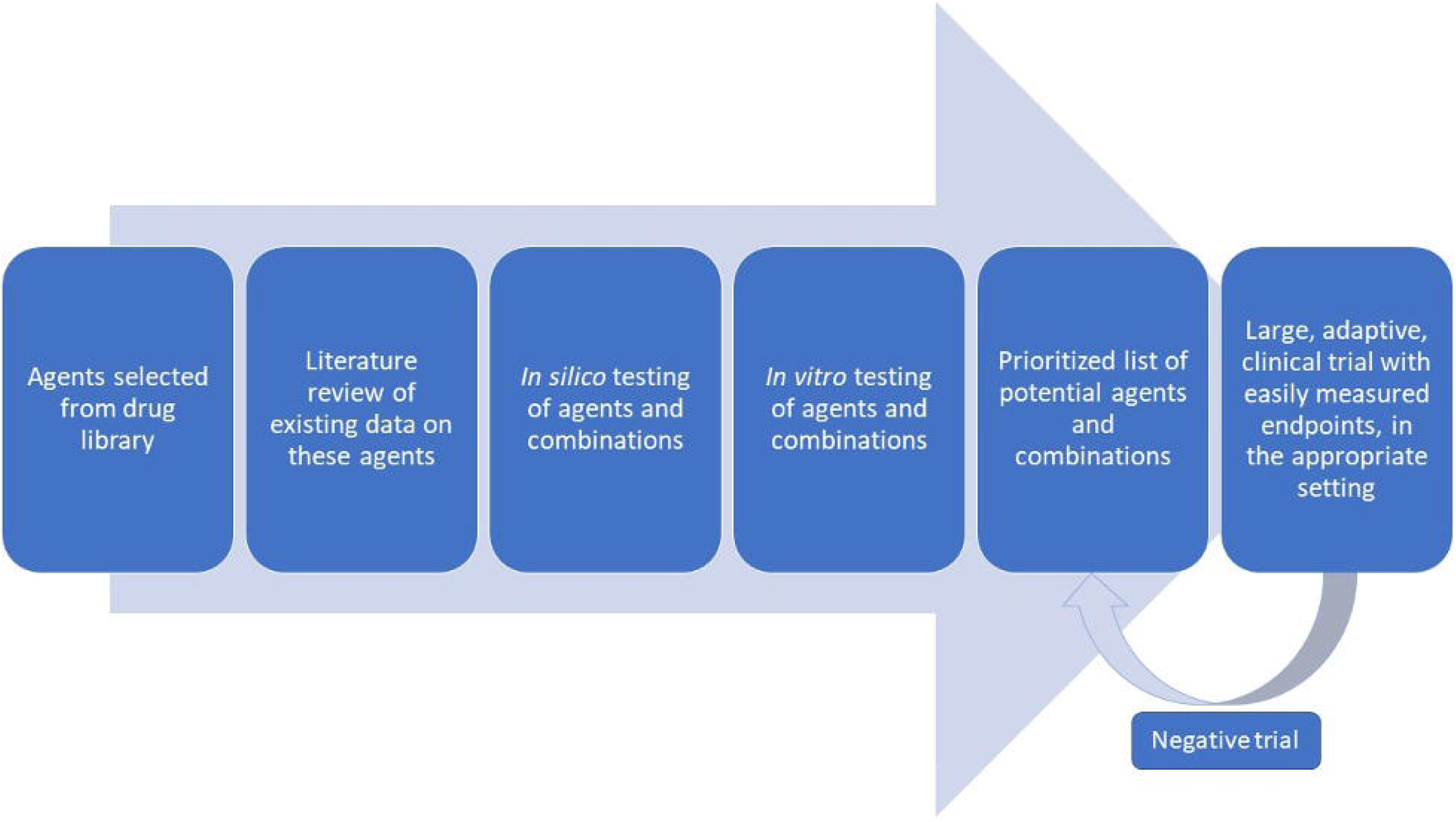
Proposed workflow for identifying effective COVID-19 therapeutics

Using a library of known compounds has the benefit not only of rapid development and established drug safety profiles, but also higher likelihood of avoiding licensing costs, making delivery to LMICs both more rapid and less costly to facilitate widespread access. Initial identification of known candidate agents for treatment of COVID-19 has mostly been achieved with computer modeling, or *in silico* testing^104^. Notable achievements on this front include protein interaction mapping^105^, which yielded both clinical and pre-clinical candidate agents as well as the screening of agents with established human safety profiles such as the ReFRAME library. This catalog of over 12,000 compounds has already been used to identify candidates for treatment of other infectious diseases such as tuberculosis and cryptosporidiosis^106^. Using this technique, Riva *et al.* reported the identification of new COVID-19 candidate agents such as apilimod^107^.

While screening of individual agents has progressed rapidly, screening for combination therapy or “cocktails” has been less robust, particularly when excluding trials using combinations including HCQ. While no agents may be highly effective alone, combinations of various agents may prove efficacious, as can be seen in HIV and hepatitis C^108, 109^. In these cases, preventing resistance to therapy through mutations is an important driver of combination therapy. Another rationale for combination therapy is targeting multiple points in regulatory circuits, which are often redundant to help maintain homeostasis. Well-designed pre-clinical studies with such targets in mind can help yield effective combination therapies^110^. Lastly, combination therapy, specifically using drug repurposing, has been a demonstrated means to address urgent needs for drugs in infectious disease when research lags behind clinical need^111^.

One early example of screening for combination therapies, though in preprint as of 20 November 2020, identified 16 synergistic combinations, but also 8 antagonistic combinations^112^. For example, synergy was found between nitazoxanide and several other compounds, and strong antagonism was found *in vitro* with the combination of remdesivir and hydroxychloroquine. While some clinical trials of “cocktail” therapy for COVID-19 are underway (interferon beta-1a with remdesivir, nitazoxanide with atazanavir, and nitazoxanide with ivermectin to name a few), *a priori* screening for candidate COVID-19 therapeutic combinations *in silico* followed by *in vitro* synergy studies could lead more rapidly to clinical trials of effective treatment of COVID-19 in LMICs and elsewhere. However, the complexities associated manufacturing and administration of combination therapies should be considered, and those with potential for affordability and availability in LMICs can be prioritized.

Despite a large number of ongoing clinical trials worldwide, only a small portion of these are adequately powered, randomized double-blinded placebo controlled clinical trials. Moreover, an even smaller fraction are currently being conducted in LMICs, likely due to several barriers. First, large clinical trials in LMICs are often require highly collaborative international partnerships. In-person site visits and collaboration, often integral to such work, has been curtailed in an effort to control the very pandemic they would seek to address. Second, diagnostic resources and capacity are more limited in LMICs compared to many higher-income countries. This limitation of available and accurate testing in LMICs could pose a threefold challenge: impeding modeling of the spread of the pandemic to facilitate well-planned trials, reducing identification and recruitment of proven cases, and further limiting diagnostic capacity for direct research use. Lastly, these interventions require trial expertise in multiple clinical settings: as treatment of severe, hospitalizing disease to reduce morbidity and mortality, as treatment of mild or moderate disease to reduce progression and transmission of disease, and as prophylactic or presumptive treatment. Indeed, targeted prophylaxis or presumptive treatment to high risk groups may be one of the more promising roles for such agents, with examples of success in LMICs with meningococcal disease^113^ and malaria^114^.

Though these challenges are significant, they are not insurmountable. New drug candidates appear rapidly and a coordinated adaptive trial design should be used. As the clinical context of this disease is global, focused clinical endpoints that do not require extensive resources can provide rigor without burdening researchers with added cost and complexity. Designing such trials requires broad expertise and thus broad collaboration. Such trial protocols can be then distributed, decreasing burdens on aspiring trialists and increasing sample sizes and sites, and thus improving power and generalizability of findings. Indeed, some or all of the above approaches have been used by groups such as RECOVERY, SOLIDARITY, and ACTT1-3, to name a few. These trials have provided some of the most actionable results to date, with positive and negative results. An example of work to coordinate such large efforts with open collaboration is the COVID-19 Clinical Research Coalition, which has called for and facilitated large RCTs in LMIC and has made research materials available for these studies^86^. Many of the agents in these trials, such as hydroxychloroquine, are now of lower interest given repeated negative findings. However, this framework of collaboration, in combination with the screening techniques discussed above, could lead more rapidly to much-needed therapies.

## Conclusions

Particularly for LMICs, injectable tools against COVID-19 - whether vaccines, or therapeutics such as remdesivir and monoclonal antibodies which are not included in this review - present challenges in production, distribution, and uptake. Agents that are accessible and easily administered, as well as combinations of such agents, can and should be emphasized in pragmatic, adaptive clinical trials specifically targeted for LMICs. Such efforts could yield not only more rapid results but also a truly global impact whereby we can prevent COVID-19 from becoming a neglected tropical disease.

## Data Availability

All data are fully available without restriction.
The data are held in public repositories: www.clinicaltrials.gov, www.clinicaltrialsregister.eu, and https://apps.who.int/trialsearch.

## Acknowledgements

We thank Richard Feachem and Kathryn Vosburg from the Pandemic Response Initiative, Institute for Global Health Sciences, University of California, UCSF, for their guidance on and review of the manuscript.

## Funding

This work was supported by funding from the Lampert Byrd Foundation.

## Co-author contact information

Daniel Maxwell, UT Southwestern Medical Center

Kelly C. Sanders, University of California, San Francisco (UCSF)

Oliver Sabot, UCSF

Ahmad Hachem, UT Southwestern Medical Center

Alejandro Llanos-Cuentas, Universidad Peruana Cayetano Heredia (UPCH)

Ally Olotu, Ifakara Health Institute

Roly Gosling, UCSF

James B. Cutrell, UT Southwestern Medical Center

## Notes

### Competing Interest Statement

The authors have declared no competing interest.

### Funding Statement

This work was supported by a grant from the Lampert Byrd Foundation (MSH, KS, and OS). http://www.lampertbyrdfoundation.org/

The funder did not have any role in the study design, data collection and analysis, decision to publish, or preparation of the manuscript.

